# The impact of the COVID-19 pandemic on adult mental health in the UK: A rapid systematic review

**DOI:** 10.1101/2021.08.23.21262469

**Authors:** Eleonore Batteux, Jo Taylor, Holly Carter

## Abstract

**Background:** There is evidence that the COVID-19 pandemic has affected the mental health of the UK population, but this needs synthesising to guide effective policy recommendations and ensure support is targeted to populations most at risk. We conducted a rapid systematic review of the evidence of the impact of COVID-19 and associated restrictions on the mental health of UK adults, including risk and protective factors.

**Method:** A range of databases were searched to identify eligible studies. Studies were eligible if they reported primary quantitative or qualitative research on the mental health of UK adults between March 2020 and March 2021. Journal publications and pre-prints were included. Reviews, position papers, protocol papers and studies published in languages other than English were excluded. The study authors screened papers for eligibility and included 102 papers in the analysis.

**Results:** The evidence from this review indicates that the mental health of UK adults has declined since the start of the pandemic, with different populations being unequally affected. Populations particularly affected are women, young adults, ethnic minorities, people from lower socio-economic backgrounds, people with pre-existing conditions and people who have had COVID-19. Other risk factors include having to isolate and time spent watching pandemic related news. Protective factors include social contact and maintaining healthy behaviours, such as physical activity.

**Conclusions:** Policy should aim to discourage risky behaviours while ensuring support is available for people to engage in protective behaviours. Interventions should be directed towards populations that have been most adversely affected. Addressing the decline in mental health across the UK population since the COVID-19 pandemic will require increasing mental health provision and ensuring equitable access to support.

## Introduction

In response to the global COVID-19 pandemic, the UK government introduced national lockdown restrictions in March 2020 [1]. People were instructed to stay at home and only permitted to leave once a day to buy essentials or take exercise. Schools and most workplaces closed, with a significant proportion of the population temporarily or permanently out of work. Since then, the UK has been through three national lockdowns, with some variation in implementation between the four nations [2]. In between national lockdowns, regions in the UK have also experienced varying levels of restrictions, with some nearly as strict as the national lockdowns themselves [3]. It is crucial that the impact of these restrictions on mental health are evaluated, with recommendations made to tackle any detrimental effects and assess the longer term needs of the population.

Many studies have been conducted on mental health during the COVID-19 pandemic, including in the UK [4,5]. These broadly suggest that mental health of the UK population has been negatively affected [6], with some people being more affected than others [7]. Where systematic reviews on the impact of the COVID-19 pandemic on mental health have been conducted, these are not specific to the UK [8, 9] and often focused on certain groups, such as healthcare professionals [10] or pregnant women [11]. A systematic review which provides a comprehensive assessment of the mental health impact of COVID-19 in the UK, alongside policy recommendations, is lacking.

To address this gap, we conducted a rapid systematic review of the impact of the COVID-19 pandemic on mental health in the UK, including general indicators of mental health outcomes, risk factors and protective factors. This allowed us to identify which populations are most affected, highlight potential interventions to protect mental health and provide a comprehensive overview of the impact of the COVID-19 pandemic.

## Methods

### Eligibility criteria

Studies were eligible if they reported primary quantitative or qualitative research on the mental health of UK adults during the COVID-19 pandemic, i.e. since March 2020. The review focused on mental health symptoms commonly experienced by the general population, which included depression, anxiety and stress, as well as self-harm and suicide. Psychiatric conditions such as psychosis and bipolar disorder were therefore excluded, as were studies on specific professional groups such as healthcare workers. Reviews, position/discussion papers, conference abstracts, protocol papers and studies published in languages other than English were excluded.

### Search

Two systematic searches were conducted. The first search took place in January 2021 for papers between 01/03/2020 and 25/01/2021 and the second in March 2021 for papers between 22/01/2021 and 15/03/2021. Sources searched included Medline, Embase, PsycINFO, iSearch Covid 19 portfolio (preprints only), Evidence Aid, Ireland National Health Library and Knowledge Service Evidence Summaries, NICE Covid guidelines, Cochrane Special Collections, Oxford Covid19 Evidence Service, CADTH, Health Information and Quality Authority Ireland, SPORE Evidence Alliance. Search terms included COVID-19 related terms (e.g. coronavirus, COVID-19, SARS-Cov2), mental health related terms (e.g. wellbeing, anxiety, stress, depression) and UK related terms (e.g. United Kingdom, NHS, Britain). A complete list of search terms is available in Supplementary File 1.

### Study identification

The study identification process is detailed in Figure 1. Records underwent initial title and abstract screening, with any relevant records then being screened by full text. The main reasons for exclusion were if papers did not report new data or data were not exclusively from the UK (which was not always obvious from the title or abstract). Some were excluded because they were preprints which duplicated later publications. By the end of this process, 102 studies were deemed eligible and included in the review. A list of included studies along with their methodological characteristics can be found in Supplementary File 2.

**Figure 1:**
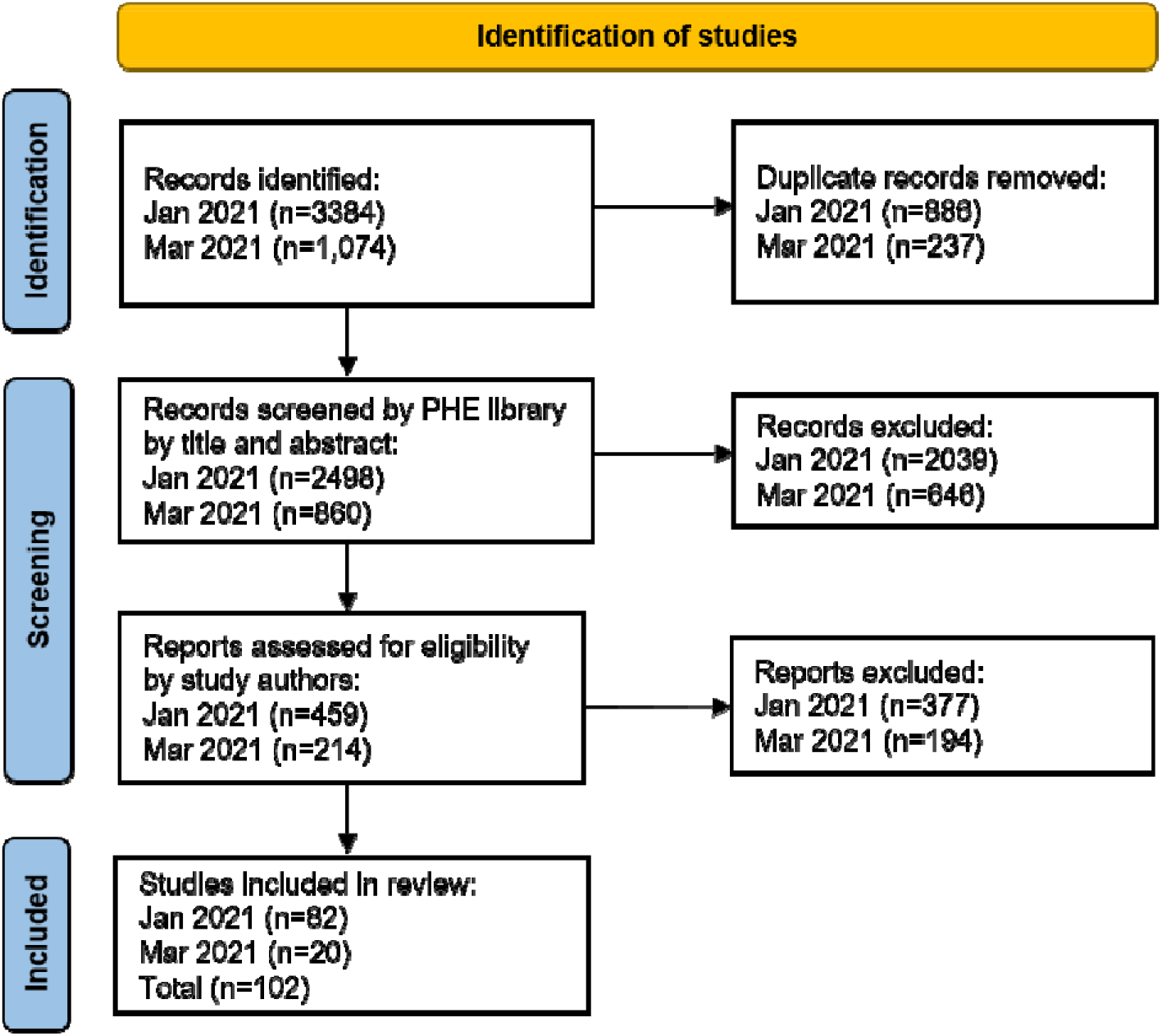
PRISMA flow diagram of the identification of studies process.

## Results

### Mental health outcomes

There is overwhelming evidence that there has been a deterioration in mental health across the UK population compared to before the COVID-19 pandemic [5, 4, 12, 13]. The decline was sharp when lockdown measures were introduced in March 2020 but reduced over time [14]. Prevalence of adverse mental health symptoms increased from 24% in 2017-2019 to 38% in April 2020 and remained elevated in May (35%) and June (32%) 2020 [15]. Around 29% of adults whose symptoms were not reflective of a mental disorder less than a year earlier were experiencing symptoms that were reflective of one by April 2020, although this has decreased over time [16].

#### Anxiety and depression

Levels of depression and anxiety during the COVID-19 pandemic have been higher than pre-pandemic levels [17, 18, 19], accompanied by increased loneliness [20] and psychological distress [13]. This seemed to be particularly the case when lockdown measures were first introduced. Although there was a sharp rise when lockdown measures were introduced, levels of depression and anxiety decreased over time and plateaued as restrictions eased, suggesting individuals adapted to the circumstances [14, 6]. Given this, one might expect that mental health services would have received more referrals, but the evidence on this is mixed. Primary care services experienced fewer than expected reported incidences of depression and anxiety disorders between April and September 2020 [21, 22, 23]. This could be due to reduced availability of GP appointments leading patients to bypass primary care or a reluctance to visit primary care services given the risk of transmission. While referrals to secondary care mental health clinical services in Cambridgeshire and Peterborough initially dropped as the first lockdown was introduced, there was a longer-term acceleration in the referral rate [24, 25]. Admissions to an acute medical unit in Bristol for patients with mental health problems increased during the first lockdown compared to 2019 [26].

#### Self-harm and suicide

Suicidal thoughts have increased [27, 28], although we do not yet know whether this has affected suicide rates. The evidence relating to rates of self-harm is mixed. In Oxford and Derby, there was a decline in hospital presentations for self-harm during the three months of the first lockdown [29], which affected women more than men. Reasons could include a reduction in self-harm at the community level, as well as individuals avoiding presenting to hospital following self-harm. COVID-19 related factors such as reduced support services and isolation were identified as influences in nearly half of self-harm presentations in Oxford and Derby, particularly in women [29]. Experiences of physical or psychological abuse during the pandemic have also been found to be associated with thoughts and instances of self-harm [30]. In Birmingham, there was an increase in presentations of self-harm between March and May 2020 compared to the previous year [31].

#### Substance use

Alcohol use has increased, whereas the evidence for smoking is mixed. Alcohol use has increased for more than one in six UK adults, associated with poor mental health [32, 4, 33]. Alcohol consumption increased in lockdown for those with pre-existing alcohol use disorders [34]. Some smokers reported an increase in smoking as a coping mechanism [35], while others reported a decline due to lockdown enabling quitting through the lifting of social barriers [4, 35].

#### Eating disorders

COVID-19-related stress and anxiety was associated with negative body image [36], body dissatisfaction and maladaptive eating behaviours [37] particularly among those with a current or past diagnosis of eating disorders [37].

### Risk factors

While there has been an increase in mental health problems across most sociodemographic groups, this increase is more pronounced for some groups than others [15].

#### Demographic risk factors

##### Gender

Women have been more affected by poor and worsening mental health since the start of the pandemic [38, 39, 40, 17, 18, 27, 4, 41, 42, 12, 13, 6, 15, 40, 43, 19, 44]. They have also experienced more physical and emotional abuse [28], thoughts of self-harm and suicide [28], loneliness [45] and increased drinking [33]. New mothers are also experiencing difficulties, with 56% reporting low mood and 71% worried, although 70% report feeling able to cope [46]. A more equal division of household chores was associated with better coping [46]. However, it is worth noting that women seem to be recovering quicker than men after worsening mental health in the initial stages of lockdown [14]. In addition, a high proportion of LGBTQ+ individuals (69%) have experienced symptoms of depression during the pandemic, with 1 in 6 reporting some form of discrimination [47]. Trans and gender diverse people have also reported a lack of access to mental health services and gender-affirming interventions [48].

##### Age

Young people have been particularly susceptible to increased anxiety and depression [40, 17, 18, 27, 4, 42, 43, 13, 6, 15, 40, 49, 44] and increased loneliness [20, 45], with students particularly at risk [50, 51]. As many as 84% of young people experienced symptoms of depression in the first 6 weeks of the first lockdown, with 72% experiencing symptoms of anxiety [43]. Younger individuals with lower incomes and previous mental health diagnoses experienced higher levels of anxiety following announcements of lockdown [52]. Some young people have decreased their drinking behaviour whereas others have increased it [33]. In addition, the mental health of older people has declined disproportionately, which is a concern given the higher propensity to be lonely, shielding and worry about death and illness [53, 54, 49].

##### Ethnicity

Minority ethnic groups have experienced a greater decline in mental health, independently of other demographic and socioeconomic characteristics [39, 4], although one study did not find a relationship between ethnicity and mental health decline [28]. There is some conflicting evidence about which minority ethnic groups have been most affected, with one study finding that young people identifying as Black/Black-British ethnicity were most at risk [55] and others that people from Asian backgrounds have been most affected [39, 4]. Minority ethnic groups have also experienced more physical and emotional abuse and thoughts of self-harm and suicide [28].

##### Socioeconomic group

People who are on lower income, financially vulnerable or worried about money have experienced higher levels of depression, anxiety, stress and loneliness [38, 56, 57, 58, 59, 18, 27, 42, 2, 6, 51, 60, 61]. This has been linked to adverse experiences such as redundancies [62, 63]. Indeed, people who are unemployed [18, 64, 41], relying on government benefits [65] and experiencing food and housing insecurity [61] all experienced worse mental health. Abuse, both mental and physical, and thoughts of self-harm and suicide have been higher in the socioeconomically disadvantaged and unemployed [7]. Thoughts and instances of self-harm are also higher in those who have had financial worries [30]. Being stressed about finances has also led to increased drinking [33] as have higher income and education [33].

##### Living conditions

Where people live and who they live with influences their mental health. Those living in urban areas [51], deprived areas [13] and without outdoor space [66] have experienced worse mental health. People living alone are particularly at risk [41, 20, 6, 45, 44], which makes sense given the association between loneliness and depressive symptoms during the pandemic [19]. People in poor partner relationships [67 (Dickerson et al, 2020) and with young children [17, 6] have also experienced worse mental health, particularly early on in the March 2020 lockdown for those caring for young children [68]. Those caring for adult children experienced a sustained increase in anxiety until July 2020 [68]. Living with children has also been linked to increased alcohol consumption [69]. Finally, people with caring responsibilities have been particularly affected [70, 71, 72], although there is some evidence that they feel like their life is more worthwhile than those without caring responsibilities [72].

##### Health conditions

A wide range of people with pre-existing physical health conditions have experienced a greater decline in mental health [59, 42, 7, 73, 74, 75, 70], including increased physical and emotional abuse and thoughts of self-harm and suicide [7]. The situation has also worsened for those with pre-existing mental health problems [27, 42, 7, 6, 51, 12] which has led to increased drinking [33]. People living with dementia have been affected due to reduced access to social support services [76, 77]. Up to 90% of children and young adults with physical and/or intellectual disabilities reported a negative impact on their mental health, often due to a lack of access to specialist facilities, therapies and equipment [78].

#### COVID-19-related risk factors

##### COVID-19 patients

People with confirmed or suspected COVID-19 were more likely to suffer from a decline in mental health [41, 4]. They experienced more loneliness [41], self-harm and suicidal thoughts [7, 30] and psychological and physical abuse from others [7]. Patients experienced higher symptoms of depression and impaired quality of life on post-hospital discharge compared to controls [79]. PTSD symptoms were also elevated in those who had been admitted to hospital with COVID-19 compared to those who were not hospitalised and experienced milder COVID-19 symptoms [80].

##### COVID-19 anxiety

COVID-19 related anxiety also led to reduced well-being and increased depression and anxiety [12, 81, 82, 83]. In some cases, this has led to acute stress presentations to hospital [84]. It seems to be higher in women, those with lower income [85, 86] and perhaps in older adults [59], although not in all studies [85].

##### At-risk groups

People at high risk of COVID-19 have experienced a decline in mental health [38, 87, 12, 65, 66, 88], with nearly half (44.5%) reporting that their mental health worsened during the first lockdown [88].

##### Isolation

Having to self-isolate [57] and shield [66, 82, 89, 90] have led to poorer mental health. Indeed, feelings of loneliness and isolation were a key risk factor for poorer mental health [53, 91; 61, 57].

##### Media

Increased time spent following COVID-19 news led to declines in mental health [51]. Employed people and students seem to be more prone to adverse reactions such as paranoia and hallucinatory experiences in response to COVID-19 news [92]. Increased screen time was also associated with poorer mental health [40].

### Protective factors

#### Social support

Social support, cohesion and connectedness were protective against depression, anxiety and loneliness [57, 20, 45, 81, 28, 58, 67]. Indeed, many spoke to friends or family to support their mental health [93]. Experiencing social contact, both face to face or via phone or video was associated with fewer depressive symptoms [94]. Greater neighbourhood identification was associated with a stronger social network and better mental health outcomes [65]. Having a pet or regular animal interaction during lockdown was also protective of good mental health [95].

#### Self-care

Other coping mechanisms included self-care activities such as mindfulness, meditation or yoga [93, 41, 49]. Others stay occupied with gaming [93].

#### Healthy behaviours

Changes to diet and sleep activity affected mental health during lockdown [69, 96]. Maintaining routine was found to be helpful among older adults [81]. People who exercised less during lockdown experienced worse mental health [32, 91], particularly those who lowered their physical activity compared to before restrictions [69]. Those who stayed active often did so to protect their mental health [19]. Outdoor activities such as exercise and gardening also improved mental health [51]. Those who spent more time outside had better sleep, which is associated with better mental health [96].

## Discussion

This systematic review draws together relevant research to provide a comprehensive overview of the impact of COVID-19 on the mental health of UK adults, while identifying risk and protective factors of poor mental health. It is intended for use by people who would benefit from an overarching picture to feed into their decision-making, such as policymakers shaping mental health services and where to direct resources to.

Findings demonstrate that the mental health of the UK adult population has declined since the start of the pandemic compared to previous years. This echoes global trends where there are many reports of increases in anxiety and depression and studies highlighting the impact of loneliness and social isolation [17]. Increases in thoughts of self-harm and suicide align with previous infectious disease-related public health emergencies, during which there tends to be a higher risk of suicidal thoughts, behaviour, and deaths [23]. In this review, we find evidence that while mental health declined sharply when lockdown was introduced in March 2020, it improved over time. This could be due to a range of factors, such as the introduction of employment support, learning to cope with lockdown, and restrictions easing. Supporting individuals in the lead-up to future lockdowns will be key to reducing distress [6].

Not all groups have been affected to the same extent. There is ample evidence that women, younger adults, ethnic minority groups, people living in financial difficulty and people with physical and mental health conditions have been worse affected. There is some evidence that LGBTQ+ individuals, older adults, people living alone and in urban areas are more at risk. There is also evidence for risk factors specific to COVID-19, such as having had COVID-19, having to shield or self-isolate, COVID-19 anxiety and following COVID-19 news.

Findings revealed that people use a range of coping mechanisms to protect their mental health. In particular, maintaining healthy routines relative to exercise, diet and sleep have been shown to be protective, as have social support and engaging in self-care activities such as meditation.

### Policy recommendations

The impact of the COVID-19 pandemic on the mental health of the UK population demonstrates the need for a clear and focused mental health strategy to tackle these effects and avoid overloading mental health services [97]. Based on the evidence presented in this review, we make the following recommendations to inform such a strategy.

#### Tackle risk factors while facilitating protective behaviours

Increased financial hardship as a result of COVID-19 restrictions has resulted in poorer mental health. Policies which provide more financial security and employment support should help to alleviate the mental health decline. Alongside this, interventions which increase access to social support and engagement in healthy behaviours such as exercise are likely to have a positive impact on mental health. This will require giving people the means to do so, such as equipment and space for people to exercise [98, 53]. Discouraging risky behaviours such as focalising on pandemic-related news and substance use as a coping mechanism may also be helpful.

#### Increase access to mental health services

There is evidence of lower access to mental health services during lockdown [23], meaning people may not have been getting the support they need. This is likely to lead to additional demand on mental health services, particularly given declines in mental health and COVID-19 patients experiencing PTSD [99, 6]. To cope with this demand, mental health services should build partnerships with charities and organisations who can provide counselling and befriending services. Increasing mental health provision will be required, allied with governance and funding to monitor the mental health situation post-pandemic [100, 56], particularly related to suicide and self-harm. A proactive approach to discharging COVID-19 patients will be needed to ensure problems are identified and appropriately managed [101, 57]. Online interventions have been recommended as they are low-cost, scalable and readily deployable solutions which can tackle cognition and mood in real time, while also allowing for personalised interventions [102, 58]. Interventions such as low-intensity CBT can be delivered effectively remotely [20]. The challenge is motivating their continuous use and ensuring those with little or no access to technology are not left behind.

#### Ensure support is accessible to all

Some have turned to professional care to support their mental health [103]. Of concern is that populations particularly affected have struggled to access mental health services, including ethnic minority groups, lower socioeconomic groups and those with mental health conditions [40, 39, 104, 44, 105]. Indeed, groups already at risk of poor mental health before the pandemic have remained at risk throughout lockdown and its aftermath [6]. Information and guidance need to be widely shared, translated in different languages and easily accessible, especially to those who do not have access to technology. They should promote the use of inclusive messaging, informal communication channels and collaboration with caregivers to engage hard to reach populations. To ensure diverse needs are met, cross-communal sharing of information to facilitate re-design and service user involvement in planning, design and delivery should be encouraged.

### Limitations and future directions

This review shows that there is a reasonable amount of evidence already available on the negative impact of the COVID-19 pandemic on mental health, including the groups that have been particularly affected. However, it does present limitations. Due to the large amount of research available on mental health since the start of the pandemic and the broad focus on the UK population as a whole, evidence relating to less common psychiatric disorders was deemed out of scope for this review. Future research should seek to address whether the COVID-19 pandemic has also increased the prevalence of conditions such as psychosis and bipolar disorder, as has been previously discussed [106]. This review was restricted to UK adults, although a systematic review of children and adolescents’ mental health alongside recommendations would be beneficial in light of restrictions such as school closures and evidence they have been impacted as well [107, 108]. The review was also limited to UK-based studies, whereas it would be useful to conduct a more international review to understand the effects of the pandemic beyond the UK and inform recommendations on a global scale.

Due to the rapid nature of this review, a risk of bias assessment of each included study was not conducted. However, we can speak to limitations across studies which limit the findings of this review. A significant proportion of studies in this review analysed data collected from two large online survey studies, namely the COVID-19 Social Study and UK Household Longitudinal study, both of which relied on self-report data. There was a limited number of studies relying on inpatient admissions and referrals to mental health services, which will be particularly important moving forward as these may increase as people become more likely to access these services out of lockdown restrictions. Most studies relied on online surveys, which limits the inclusion of those who do not use or do not have access to the internet. This could introduce bias in responses, particularly given those without access to the internet due to financial difficulty may be more at risk of poor mental health [57]. To minimise this bias, future studies should aim to gather data via other means, such as telephone or post.

There are areas related to mental health that this review cannot address due to a lack of available evidence. Future research should address potential anxieties related to the easing of restrictions, such as socialising with others and returning to the workplace. Dealing with a changing employment landscape and any impact on the economy might also present challenges. Some may continue to experience COVID-19 anxiety despite lower prevalence of COVID-19 or having been vaccinated. The long-term impact of post-acute COVID-19 on mental health will also need to be monitored. In the UK, NHS Improving Access to Psychological Therapies (IAPT) services are already working closely with long COVID-19 assessment clinics to provide mental health support for those with the condition [109], although the efficacy of this strategy ought to be evaluated as further support may be required.

The vast majority of studies included in this review focused on data collected between March and August 2020, thereby largely reflecting the impact of the first lockdown. Research is needed on the impact of subsequent restrictions and lockdowns in the UK. It is possible that people have been able to adjust to living under these conditions, or alternatively that mental health has worsened to a greater extent given repeated lockdowns. Recent evidence indeed suggests that symptoms of depression have increased between January and March 2021 since November 2020 [110]. Finally, future research should investigate the longer-term effects of the pandemic to identify those who require further support once restrictions are lifted, which will inform mental health strategies for future pandemics.

## Data Availability

The data extracted from the studies reviewed are included in the article and Supplementary File 2.

## Availability of data and materials

Data sharing is not applicable to this article as no datasets were generated or analysed during the current study.

## Ethical statement

Not applicable.

## Author contributions

EB and JT conducted the systematic review and wrote the manuscript. HC supervised the conduct of the review and contributed to writing the manuscript. All authors read and approved the final manuscript.

## Acknowledgments

We thank the PHE Library Services for their support with the study search and identification process.

