## Supplementary File 1 for "The impact of the COVID-19 pandemic on adult mental health in the UK: A rapid systematic review"

Supplementary File: Search terms used

**Terms used:**

exp Coronavirus Infections/ OR ((corona* or corono*) adj1 (virus* or viral* or virinae*)) OR (coronavirus* or coronovirus* or coronavirinae or CoV or HCov*) OR ("2019-nCoV" or 2019nCoV or nCoV2019 or "nCoV-2019" or "COVID-19" or COVID19 or "CORVID-19" or CORVID19 or "WN-CoV" or WNCoV or "HCoV-19" or HCoV19 or "2019 novel*" or Ncov or "n-cov" or "SARS-CoV-2" or "SARSCoV-2" or "SARSCoV2" or "SARS-CoV2" or SARSCov19 or "SARS-Cov19" or "SARSCov-19" or "SARS-Cov-19" or Ncovor or Ncorona* or Ncorono* or NcovWuhan* or NcovHubei* or NcovChina* or NcovChinese* or SARS2 or "SARS-2" or SARScoronavirus2 or "SARS-coronavirus-2" or "SARScoronavirus 2" or "SARS coronavirus2" or SARScoronovirus2 or "SARS-coronovirus-2" or "SARScoronovirus 2" or "SARS coronovirus2")

AND

(Anxiety OR psychological* or psychiatric* OR stress OR depression or depressive OR Stress, Psychological/ OR Adaptation, Psychological/ or exp *Emotions/ OR (mental* adj2 (problem* or disorder* or ill* or well-being or wellbeing)) OR (mental* adj2 health) OR (SMI or psycho* or schizo* or manic or mania or bipolar or antipsycho*) OR (phobi* or agoraphobi* or anxious or obsess* or compulsi* or panic) OR ((personality or character) adj3 disorder*) OR (self-harm or self-injury or self harm or self injury)

AND
(exp United Kingdom/ OR national health service* or nhs* OR (english not ((published or publication* or translat* or written or language* or speak* or literature or citation*) adj5 english)).ti,ab. (95913)

(gb or "g.b." or britain* or (british* not "british columbia") or uk or "u.k." or united kingdom* or (england* not "new england") or northern ireland* or northern irish* or scotland* or scottish* or ((wales or "south wales") not "new south wales") or welsh*)
